# Automated differentiation of malignant and benign primary solid liver lesions on MRI: an externally validated radiomics model

**DOI:** 10.1101/2021.08.10.21261827

**Authors:** Martijn P.A. Starmans, Razvan L. Miclea, Valerie Vilgrain, Maxime Ronot, Yvonne Purcell, Jef Verbeek, Wiro J. Niessen, Jan N.M. Ijzermans, Rob A. de Man, Michail Doukas, Stefan Klein, Maarten G. Thomeer

**Author notes:** **Contact information corresponding author:** Martijn P. A. Starmans, Mail address, Address: Erasmus MC, P.O. box 2040, 3000 CA, Rotterdam, The Netherlands. equal contributions (Stefan Klein, Maarten G. Thomeer). **Mail:**. **Data availability statement:** Imaging and clinical research data are not available at this time. Programming code is available on Zenodo at DOI https://doi.org/10.5281/zenodo.5175705. **Financial support statement:** No funding sources were involved in the study design, collection, analysis, and interpretation of data, writing the report, nor the decision to submit the article for publication. **Author Contributions:** M.P.A.S., R.L.M., V.V., M.R., W.J.N., S.K. and M.G.T. provided the conception and design of the study. M.P.A.S., R.L.M., Y.P., J.V., J.I., R.A.d.M., M.D. and M.G.T. acquired the data. M.P.A.S., S.K. and M.G.T. analyzed and interpreted the data. M.P.A.S. created the software. M.P.A.S., S.K. and M.G.T. drafted the article. All authors read and approved the final manuscript.

## Abstract

**Background & Aims:** Distinguishing malignant from benign primary solid liver lesions is highly important for treatment planning. However, diagnosis on radiological imaging is challenging. In this study, we developed a radiomics model based on magnetic resonance imaging (MRI) to distinguish the most common malignant and benign primary solid liver lesions, and externally validated the model in two centers.

**Approach & Results:** Datasets were retrospectively collected from three tertiary referral centers (A, B and C) including data from affiliated hospitals sent for revision. Patients with malignant (hepatocellular carcinoma and intrahepatic cholangiocarcinoma) and benign (hepatocellular adenoma and focal nodular hyperplasia) lesions were included. For each patient, only a T2-weighted MRI was included. A radiomics model was developed on dataset A using a combination of machine learning approaches, and internally evaluated on dataset A through cross-validation. Next, the model was externally validated on datasets B and C, and compared to scoring by two experienced abdominal radiologists on dataset C. In the resulting dataset, in total, 486 patients were included (A: 187, B: 98 and C: 201). Despite substantial MRI acquisition heterogeneity, the radiomics model developed on dataset A had a mean area under the receiver operating characteristic curve (AUC) of 0.78 in the internal validation on dataset A, and a similar AUC in the external validations (B: 0.74, C: 0.76). In dataset C, the two radiologists showed moderate agreement (Cohen’s κ: 0.61) and achieved AUCs of 0.86 and 0.82, respectively.

**Conclusions:** Our radiomics model using T2-weighted MRI only can non-invasively distinguish malignant from benign primary solid liver lesions. External validation indicated that our model is generalizable despite substantial differences in the acquisition protocols.

## Introduction

Liver cancer is the seventh most commonly diagnosed cancer and the third most common cause of cancer deaths worldwide, with approximately 906,000 estimated new cases and 830,000 deaths in 2020 (1). One of the most important tasks in routine clinical practice is making the distinction between malignant and benign primary solid liver lesions, which substantially influences treatment planning (2, 3). Commonly, a first assessment is made by the radiologist based on imaging, generally magnetic resonance imaging (MRI). Guidelines such as those from the European Association for the Study of the Liver (EASL) (4, 5) may aid the radiologist. Typically, a mixture of T2-weighted, T1-weighted, dynamic contrast enhanced MRI, diffusion weighted imaging, and the apparent diffusion coefficient (ADC) is used. The diagnosis is often challenging due to the wide variety of liver lesion phenotypes, sizes, and appearances (6), and lack of a clear assessment consensus (7).

Patients from peripheral centers may therefore be referred to tertiary centers for reassessment. This trajectory is time consuming and expensive, while a quick and accurate diagnosis is crucial for the treatment planning. Often, despite imaging, a biopsy may be performed to make the final diagnosis, as indicated by the EASL guidelines. While accurate, biopsies are (minimally) invasive, can be technically challenging, and bring risks such as bleeding and tumor seeding to the patient (8). Patient treatment may benefit from a non-invasive tool to shorten time to diagnosis by enabling quicker referral, refining patient selection prior to biopsies, and assist diagnosing patients who do not require a biopsy.

In recent years, radiomics, i.e., the use of a large number of quantitative medical imaging features to predict clinical outcomes, has been successfully used in various clinical areas (9-11). In liver cancer, this has been mostly based on computed tomography to make predictions such as survival, prognosis, and recurrence (12-14). For MRI in liver cancer, radiomics has been used to classify focal liver lesions (15-18), and as LI-RADS (19) surrogate (20). Radiomics thus shows potential for usage in liver lesion characterization.

However, as concluded in a recent review, the use of radiomics for liver lesion characterization is still at an early stage (21). First, there is a need for large, multicenter cohorts, especially for external validation (22-24). Second, a major challenge is the lack of image acquisition standardization (21), as radiomics methods are generally sensitive to acquisition variations (25), underlining the need for external validation. Rather than requiring a comprehensive, standardized set of multiple MRI sequences, usage of a single sequence would make radiomics models more universally applicable in a routine clinical setting.

The primary aim of this study was therefore to develop a radiomics model based on only T2-weighted MRI to distinguish between the most common malignant and benign primary solid liver lesions, and to externally validate the model in two multicenter cohorts. We used only T2-weighted MRI, as this sequence is widely available, reliable for lesion segmentation, minimally sensitive to motion or breathing artefacts, and informative (4, 5, 19). Our secondary aim was to compare the performance of radiomics to clinical practice through visual scoring of the lesions by two experienced abdominal radiologists.

## Materials and Methods

### Data collection

Approval for this study by the institutional review boards of Erasmus MC (Rotterdam, the Netherlands) (MEC-2017-1035), Maastricht UMC+ (Maastricht, the Netherlands) (METC 2018-0742), and Hôpital Beaujon (Paris, France) (N° 2018-002) was obtained. Informed consent was waived due to the use of retrospective, anonymized data. The study protocol conformed to the ethical guidelines of the 1975 Declaration of Helsinki.

Three datasets were collected retrospectively from three tertiary referral centers: all patients diagnosed or referred to A) Erasmus MC between 2002 - 2018; B) Maastricht UMC+ between 2005 - 2018; and C) Hôpital Beaujon, included in reverse chronological order starting at 2018, until in total 201 patients were identified, in accordance with the inclusion and exclusion criteria described below. Imaging data, age, sex, and phenotype were collected for each patient.

Inclusion criteria were: hepatocellular carcinoma (HCC), intrahepatic cholangiocarcinoma (iCCA), hepatocellular adenoma (HCA) or focal nodular hyperplasia (FNH); pathologically proven phenotype, except for “typical” FNH; and availability of a T2-weighted MRI scan. Exclusion criteria were: maximum diameter equal to or smaller than 3 cm; underlying liver disease; and significant imaging artefacts. Details on the pathological examination are given in **Supplementary Material 1**.

Malignant lesions included HCC (75 - 85% of primary liver cancers), and iCCA (10 - 15% of primary liver cancers) (6). Benign lesions included HCA (3-4 cases per 100,000 person-years in Europe and North America) and FNH (found in 0.8% of all adult autopsies) (6). The most common benign primary liver lesions, hemangioma, were not included as these are nonsolid and often relatively easy to diagnose on imaging (4, 26). Only lesions with a pathologically proven phenotype were included to ensure an objective ground truth. Pathological analysis for each patient was performed locally in their admission hospital. An exception was made for typical FNH (6), which are routinely not biopsied and diagnosed radiologically (27), as typical FNH imaging characteristics are 100% specific (4). Not including these would create a selection bias towards “atypical” FNH: the model performance would than only be evaluated on atypical FNH, and no claims could be made on the performance in typical FNH. In patients with multiple lesions, only the largest one was included.

Patients with underlying liver disease due to alcohol, hepatitis, and vascular liver disease, such as fibrosis or cirrhosis, were excluded, as the *a priori* chance of a lesion being HCC in these patients is by far the largest (28). Steatosis was not an exclusion criterium. Diagnosis of liver disease was based on clinical, pathological and/or imaging findings. In case of HCC, cirrhosis was always excluded from biopsy or resection. Lesions with a maximum diameter equal to or smaller than 3 cm were excluded, since in non-cirrhotic livers these have a high probability of being secondary lesions, hemangioma, or cysts (26, 29), which are generally easy to diagnose on imaging (4, 26). Hence, a radiomics model would have relatively little added value in these patients with underlying liver disease or small lesions. When T2-weighted MRI with fat saturation was not available, regular T2-weighted MRI was used, similar to clinical practice. Images with significant artefacts (i.e., patient or scanner related) and therefore not suitable for diagnostic purposes, as judged by an experienced radiologist (21 years of experience), were excluded.

### Segmentation

Lesion segmentation was done semi-automatically using in-house software (15). Each lesion was segmented by one of three observers: a radiology resident, and two experienced abdominal radiologists (21 and 8 years of experience). The observers were aware of the inclusion and exclusion criteria, and were asked to segment a primary liver lesion. When the lesions could not be found, e.g. isointense lesions, the observers were able to look at the other sequences if available. The observers could segment manually or semi-automatically using region-growing or slice-to-slice contour propagation. Segmentation was performed per slice in the 2D transverse plane, resulting in a 3D volume. Semi-automatic results were always reviewed and manually corrected when necessary, to assure the result resembled manual segmentation. All segmentations were verified by the most experienced radiologist. A subset of 60 lesions (30 from dataset B, 30 from dataset C) was segmented by two observers to assess the intra-observer variability using the pairwise Dice Similarity Coefficient (DSC), with DSC > 0.70 indicating good agreement (30).

### Radiomics

An overview of the radiomics methodology is depicted in **Figure 1**. As T2-weighted MRI scans do not have a fixed unit and scale, the full images were normalized using z-scoring. No further preprocessing was performed. For each lesion, 564 features quantifying intensity, shape and texture were extracted from the T2-weighted MRI scan. For details, see **Supplementary Material 2**. To create a decision model from the features, the Workflow for Optimal Radiomics Classification (WORC) toolbox was used (31, 32). In WORC, decision model creation consists of several steps, e.g. feature selection, resampling, and machine learning. WORC performs an automated search amongst a variety of algorithms for each step and determines which combination maximizes the prediction performance on the training dataset. For details, see **Supplementary Material 3**. The code for the feature extraction and model creation has been published open-source (33).

**Figure 1.**
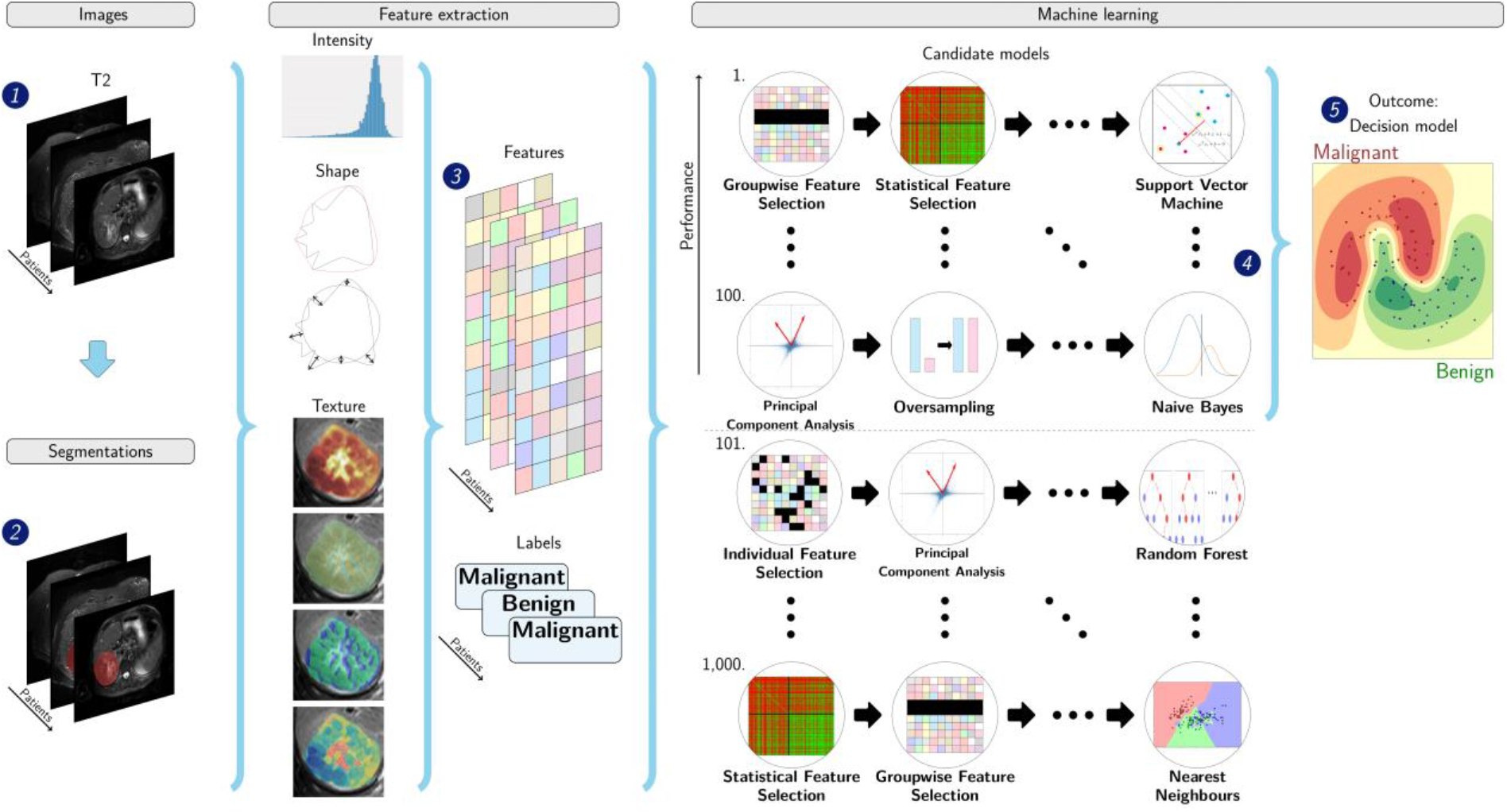
Schematic overview of the radiomics approach. Adapted from (46). Input to the algorithm are the T2-weighted MRI scans (1) and the lesion segmentations (2). Processing steps include feature extraction (3) and the creation of a machine learning decision model (5), using an ensemble of the best 100 workflows from 1,000 candidate workflows (4), where the workflows are different combinations of the different analysis steps (e.g. the classifier used).

### Experimental setup

First, to evaluate the predictive value of radiomics within a single center, an internal validation was performed in dataset A through a 100x random-split cross-validation (34, 35), see **Supplementary Figure S1 A**. In each iteration, the data was randomly split into 80% for training and 20% for testing in a stratified manner, to make sure the distribution of classes in all datasets was similar to that in the full dataset.

Second, to evaluate whether a model developed on data from one center generalizes well to unseen data from other centers, two external validations were performed by training a model on dataset A, and testing it on the unseen datasets B and C, see **Supplementary Figure S1 B**.

Third, as clinicians frequently use age and sex in their decision making, two additional models were externally validated based on: 1) age and sex; and 2) age, sex, and radiomics features.

For both the internal and external validations, model optimization was performed within the training dataset using an internal 5x random-split cross-validation, see **Supplementary Figure S1**. Hence, all optimization was done on the training dataset to eliminate any risk of overfitting on the test dataset.

### Performance of the radiologists

To compare the models with clinical practice, the T2-weighted MRI scans were scored by two experienced abdominal radiologists. They were blinded to the diagnosis, but aware of the inclusion and exclusion criteria. Classification of malignancy was made on a four-point scale to indicate the radiologists’ certainty: 1=benign, certain; 2=benign, uncertain; 3=malignant, uncertain; and 4=malignant, certain. To obtain binary scores, 1 and 2 were converted to benign, 3 and 4 to malignant. Several characteristics used in the decision making were also scored by the radiologists: presence of 1) central scar (6); 2) liquid; 3) atoll sign (36); and 4) degree of heterogeneity (scale 1-4 similar to malignancy). As the radiologists were from centers A and B, scoring was done on dataset C to prevent them from having seen the data previously.

### Statistical analysis

To evaluate the difference in clinical characteristics and explore the predictive value of the individual radiomics features between the malignant and benign lesions, per dataset, univariate statistical testing was performed using a Mann-Whitney U test for continuous variables and a Chi-square test for categorical variables. For the clinical characteristics, the statistical significance of the difference between datasets was assessed using a Kruskal-Wallis test for continuous variables, and a Chi-square test for discrete variables. P-values of the clinical characteristics were not corrected for multiple testing as these are purely descriptive: p-values of the radiomics features were corrected using the Bonferroni correction (i.e., multiplying the p-values by the number of tests).

For all models, the Area Under the Curve (AUC) of the Receiver Operating Characteristic (ROC) curve, Accuracy, Sensitivity, and Specificity were calculated. ROC confidence bands were constructed using fixed-width bands (37). The positive class was defined as the malignant lesions.

For the internally validated model, 95% confidence intervals of the performance metrics were constructed using the corrected resampled t-test, thereby taking into account that the samples in the cross-validation splits are not statistically independent (35). For the externally validated model, 95% confidence intervals were constructed using 1,000x bootstrap resampling of the test dataset and the standard method for normal distributions ((38) table 6, method 1), see **Supplementary Figure S1 B**.

For binary scores, the agreement between radiologists was evaluated using Cohen’s κ (39). For ordinal scores, i.e., degree of heterogeneity and malignancy, the correlation was evaluated using Pearson correlation (40). The AUCs of the radiomics model and the radiologists were compared using the DeLong test (41), and confusion matrices were used to analyze the agreement.

To gain insight into the radiomics model’s decision making, lesions were ranked based on the probability of a lesion being malignant as predicted by the model. Ranking was done as archetypal benign (ground truth benign, probability near 0%) - pitfall malignant (ground truth malignant, probability near 0%) - borderline (probability around 50%) - pitfall benign (ground truth benign, probability near 100%) - archetypal malignant (ground truth malignant, probability near 100%). This was done on dataset C to enable comparison with the radiologists.

For all statistical tests, p-values below 0.05 were considered statistically significant.

## Results

### Datasets

In total, 486 patients were included (A: 187; B: 98; C: 201). The clinical and imaging characteristics are reported in **Table 1**. As all centers serve as tertiary referral centers, the datasets originated from 159 different scanners (A: 52; B: 21; C: 86), resulting in substantial heterogeneity in the MRI acquisition protocols. Statistically significant differences between datasets A, B, and C included magnetic field strength (p=0.001), manufacturer (p=10^−4^), slice thickness (p=10^−32^), repetition time (p=0.006), flip angle (p=0.05), and use of fat saturation (p=10^−17^).

**Table 1.** Clinical and imaging characteristics of the datasets. The number of patients (N) in each dataset is indicated in the column header. Per dataset, the statistical significance of the difference between the malignant and benign lesions was assessed using a Mann-Whitney U test for continuous variables, and a Chi-square test for discrete variables. The statistical significance of the difference between datasets was assessed using a Kruskal-Wallis test for continuous variables, and a Chi-square test for discrete variables. Statistically significant p-values are displayed in **bold**.

| Dataset | A: Erasmus MC<br>(N=187) |  |  | B: Maastricht UMC+<br>(N=98) |  |  | C: Beaujon APHP<br>(N=201) |  |  | P |
| --- | --- | --- | --- | --- | --- | --- | --- | --- | --- | --- |
|  | Benign | Malignant | P | Benign | Malignant | P | Benign | Malignant | P |  |
| Patients | 93 | 94 |  | 55 | 43 |  | 117 | 84 |  |  |
| Age in years<sup>+</sup> | 37 [30-46] | 62 [25-70] | **10<sup>-19</sup>** | 38 [31-45] | 64 [60-71] | **10<sup>-14</sup>** | 38 [31-45] | 63 [53-68] | **10<sup>-20</sup>** | 0.69 |
| Sex |  |  | **10<sup>-12</sup>** |  |  | **10<sup>-6</sup>** |  |  | **10<sup>-17</sup>** | 0.22 |
| Male | 4 | 48 |  | 3 | 20 |  | 11 | 55 |  |  |
| Female | 89 | 46 |  | 52 | 23 |  | 106 | 29 |  |  |
| Phenotype |  |  |  |  |  |  |  |  |  | **0.003** |
| HCC |  | 81 |  |  | 28 |  |  | 47 |  |  |
| iCCA |  | 13 |  |  | 15 |  |  | 37 |  |  |
| HCA | 48 |  |  | 26 |  |  | 65 |  |  |  |
| FNH | 45 |  |  | 29 |  |  | 52 |  |  |  |
| Size |  |  |  |  |  |  |  |  |  |  |
| Imaging |  |  |  |  |  |  |  |  |  |  |
| Magnetic field strength |  |  | 0.10 |  |  | 0.45 |  |  | **0.003** | **0.002** |
| 1.0 Tesla | 1 | 4 |  | 2 | 4 |  | 1 | 3 |  |  |
| 1.5 Tesla | 76 | 82 |  | 48 | 39 |  | 74 | 68 |  |  |
| 3.0 Tesla | 16 | 8 |  | 5 | 0 |  | 42 | 13 |  |  |
| Scanner |  |  | **10<sup>-8</sup>** |  |  | **0.03** |  |  | 0.77 | **10<sup>-15</sup>** |
| Manufacturer |  |  |  |  |  |  |  |  |  |  |
| Siemens | 13 | 32 |  | 21 | 7 |  | 23 | 17 |  |  |
| Philips | 16 | 38 |  | 34 | 36 |  | 62 | 40 |  |  |
| GE | 64 | 24 |  | 0 | 0 |  | 30 | 24 |  |  |
| Toshiba | 0 | 0 |  | 0 | 0 |  | 2 | 3 |  |  |
| Slice thickness (mm)* | 6.0 - 8.0 | 6.0 - 7.0 | 0.12 | 5.0 - 6.0 | 5.0 - 5.0 | 0.08 | 5.0 - 6.0 | 5.0 - 6.0 | 0.41 | **10<sup>-32</sup>** |
| Pixel spacing (mm)* | 0.72 - 0.94 | 0.73 - 1.19 | **0.005** | 0.77 - 1.38 | 0.77 - 0.99 | 0.13 | 0.74 - 1.0 | 0.75 - 1.07 | 0.13 | 0.07 |
| Repetition time (ms)* | 1348 - 8571 | 1218 - 4844 | **0.001** | 1100 - 2805 | 1600 - 2961 | **0.007** | 1200 - 3884 | 1512 - 6058 | 0.14 | **0.006** |
| Echo time (ms)* | 89 - 100 | 80 - 100 | **10<sup>-3</sup>** | 80 - 112 | 80 - 90 | **0.04** | 80 - 120 | 80 - 103 | 0.13 | 0.62 |
| Flip angle (degree)* | 90 - 150 | 90 - 150 | 0.47 | 90 - 141 | 90 - 90 | **0.01** | 90 - 140 | 90 - 134 | 0.33 | **0.07** |
| Fat Saturation yes/no | 72/21 | 59 / 35 | **0.04** | 35/20 | 39/4 | **0.004** | 98/19 | 59/25 | **0.03** | **10<sup>-18</sup>** |

Abbreviations: GE: General Electric; HCC: hepatocellular carcinoma; iCCA: intrahepatic cholangiocarcinoma; HCA: hepatocellular adenoma; FNH: focal nodular hyperplasia; Max: maximum; P: p-value of Mann-Whitney U test for continuous variables, Chi-square for categorical variables.
<sup>+</sup>: median [Quartile 1 - Quartile 3]
\*: Quartile 1 - Quartile 3

On the subset that was segmented by two observers, the mean ± standard deviation of DSC indicated good agreement (B: 0.80±0.21; C: 0.81±0.11).

### Radiomics

The results of the radiomics model are depicted in **Table 2**. The internal validation on dataset A had a mean AUC of 0.78; the two external validations yielded a similar performance (B: 0.74; C: 0.76). The ROC curves (**Figure 2**) illustrate that the model trained on dataset A performed similar in each of the three centers.

**Table 2.** Performance of the radiomics model and the radiologists three datasets (A, B, and C). For the radiomics model, the mean (internal cross-validation) or point estimate (external validation) and 95% confidence intervals are reported.

| Evaluation | Internal cross-validation | External validation |  | Radiologist 1 | Radiologist 2 |
| --- | --- | --- | --- | --- | --- |
| Train set | A* | A | A | - | - |
| Test set | A* | B | C | C | C |
| AUC | 0.78 [0.70, 0.85] | 0.74 [0.65, 0.84] | 0.76 [0.70, 0.83] | 0.86 | 0.83 |
| Accuracy | 0.69 [0.62, 0.76] | 0.64 [0.54, 0.74] | 0.69 [0.62, 0.75] | 0.80 | 0.77 |
| Sensitivity | 0.70 [0.57, 0.82] | 0.79 [0.67, 0.91] | 0.82 [0.74, 0.91] | 0.88 | 0.87 |
| Specificity | 0.68 [0.59, 0.78] | 0.53 [0.40, 0.66] | 0.59 [0.50, 0.68] | 0.74 | 0.69 |

Abbreviations: AUC: area under the receiver operating characteristic curve.
\*Training and testing within a single dataset was done through a 100x random-split cross-validation.

**Figure 2.**
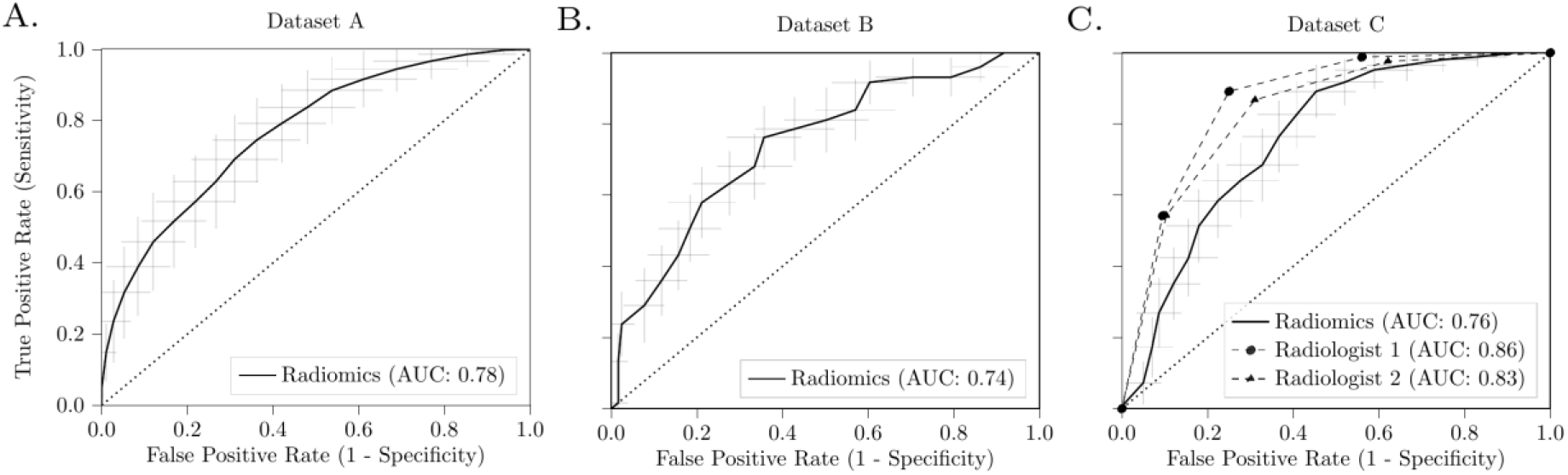
Receiver operating characteristic (ROC) curves of the radiomics model and radiologists. For the radiomics model, the curves present the model internally validated on dataset A (A); and trained on dataset A, externally validated on dataset B (B) and dataset C (C). The performance of scoring by the two experienced abdominal radiologists on dataset C is also depicted in (C). For the radiomics model, the crosses identify the 95% confidence intervals of the 100x random-split cross-validation (A) or 1,000x bootstrap resampling (B and C); the bold curves are fit through the means.

The age-and-sex-only model had a high AUC in both the internal validation (A: 0.88) and the two external validations (B: 0.93; C: 0.85). Combining age, sex, and the radiomics features yielded an improvement (A: 0.93; B: 0.98; C: 0.91), although not statistically significant. The Accuracy for the age-and-sex-only model (A:0.83; B: 0.92; C: 0.82) and the combined age, sex, and radiomics model (A: 0.85; B: 0.92; C: 0.83) were similar.

### Comparison with radiologists

The performance of the two experienced abdominal radiologists on classifying dataset C is depicted in **Table 2**. The ROC curves (**Figure 2c**) were mostly just above the 95% confidence interval of the radiomics model. The AUC of Radiologist 1 (0.87) was statistically significantly better than the radiomics model (DeLong: p=0.0028): the differences in AUC between Radiologist 2 (0.83) and the radiomics model and between the two radiologists were not statistically significant. The Accuracy per phenotype is depicted in **Table 3**. The radiomics model had a similar Accuracy in HCC (0.83) and iCCA (0.82), while the performance in FNH (0.66) was slightly better than in HCA (0.54).

**Table 3.**
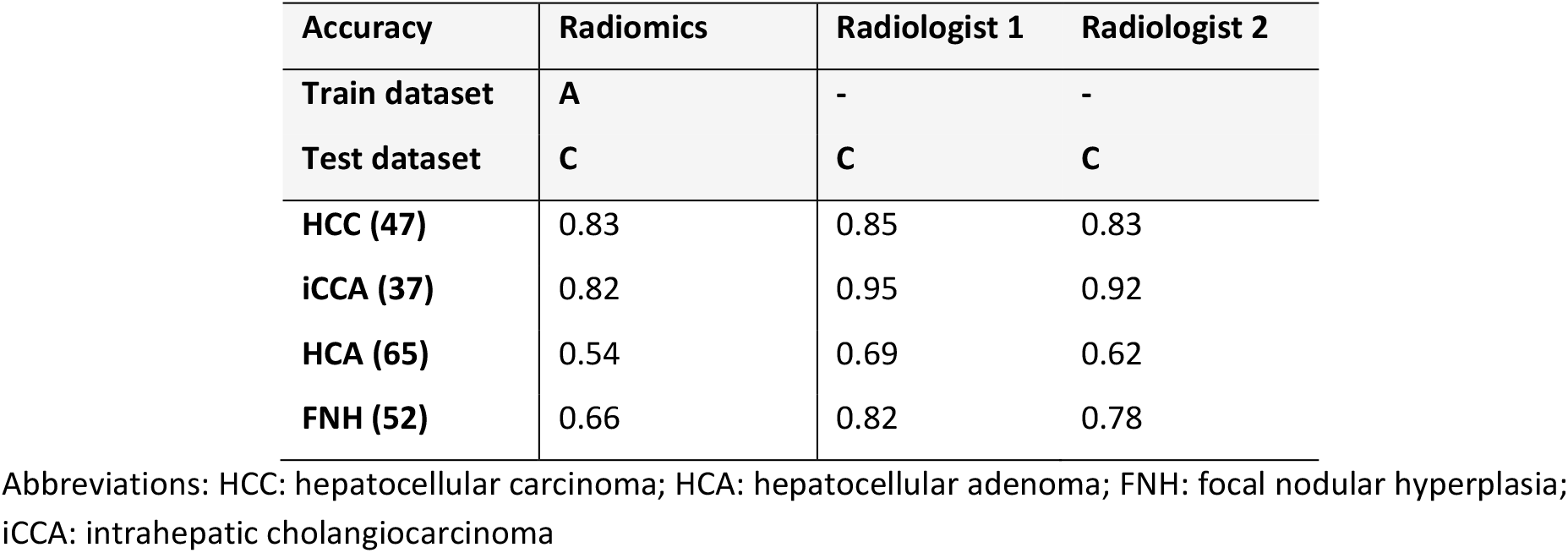
Accuracy per phenotype of the radiologists and the radiomics model in the external validation on dataset C. The Accuracy per phenotype represents the percentage of the lesions with that specific phenotype being correctly classified as malignant or benign. The number of lesions per phenotype in dataset C is given between brackets in the first column.

| Accuracy | Radiomics | Radiologist 1 | Radiologist 2 |
| --- | --- | --- | --- |
| Train dataset | A | - | - |
| Test dataset | C | C | C |
| HCC (47) | 0.83 | 0.85 | 0.83 |
| iCCA (37) | 0.82 | 0.95 | 0.92 |
| HCA (65) | 0.54 | 0.69 | 0.62 |
| FNH (52) | 0.66 | 0.82 | 0.78 |

Abbreviations: HCC: hepatocellular carcinoma; HCA: hepatocellular adenoma; FNH: focal nodular hyperplasia; iCCA: intrahepatic cholangiocarcinoma

Confusion matrices of the predictions on dataset C are depicted in **Figure 3**. The agreement between the radiologists on classifying the lesions as malignant or benign was moderate (Cohen’s κ: 0.61) (39): the two radiologists agreed in 160 of the 201 patients (80%). The agreement between the two radiologists and the radiomics model was weak (Radiologist 1: κ of 0.47; Radiologist 2: κ of 0.42), as reflected by the confusion matrices. For the other characteristics scored by the two radiologists, the agreement was weak for presence of a scar (κ: 0.41) and liquid (κ: 0.52), and strong for presence of the atoll sign (κ: 0.80); the correlation was moderate for heterogeneity (Pearson coefficient: 0.69) and strong for malignancy (Pearson coefficient: 0.70) (40).

**Figure 3.**
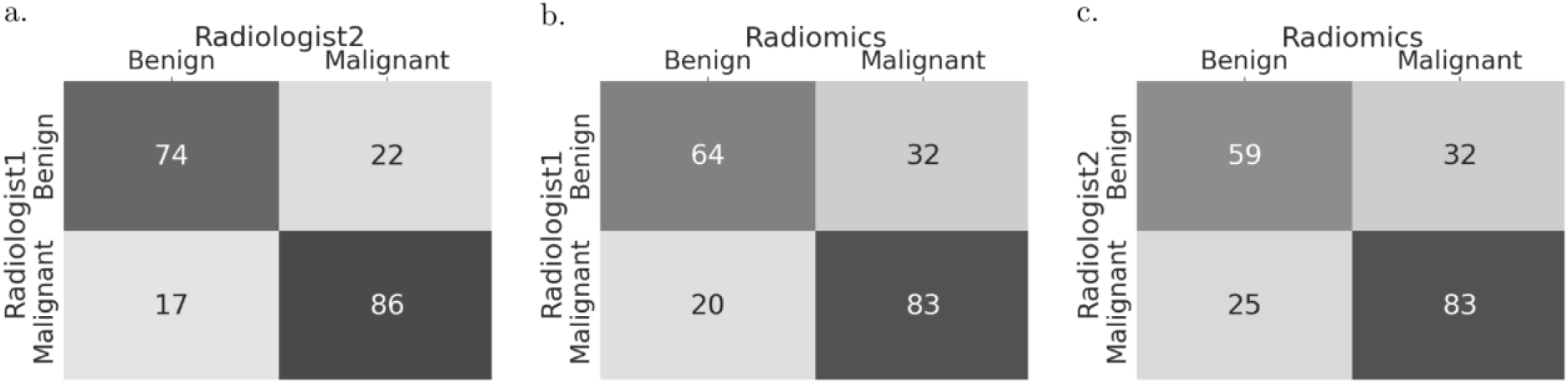
Confusion matrices of the predictions by the radiomics model and the two radiologists. The darker the background, the higher the agreement.

### Model insight

In dataset A, on which the radiomics model was developed, 45 radiomics features showed statistically significant differences between the malignant and benign lesions with p-values after Bonferroni correction from 9×10^−10^ to 0.049. These included 4 shape features (volume was not significant), 1 orientation feature, and 40 texture features. Statistically significant differences were found for 49 radiomics features in dataset B and 10 in dataset C. Four radiomics features (all texture) showed statistically significant differences in all three datasets. A list of these features and their p-values can be found in **Supplementary Table S1**. The differences in volume between the three datasets was statistically significant (p=10^−10^).

Examples of lesions from dataset C ranked as archetypal, borderline, or pitfall by the radiomics model are depicted in **Figure 4**. Visual inspection of the T2-weighted MRI scans of the archetypal or pitfall lesions showed a relation with heterogeneity (archetypal malignant: heterogeneous; archetypal benign: homogeneous), area and volume (archetypal malignant: generally high maximum axial area and high volume), and irregularity of shape on 2-D axial slices (archetypal malignant lesions: irregular; archetypal benign: compact). Pitfall lesions showed the opposite, e.g. pitfall benign: heterogeneous. Borderline lesions, i.e., with an almost equal predicted chance of being malignant or benign, were mostly of medium size and medium heterogeneity.

**Figure 4.**
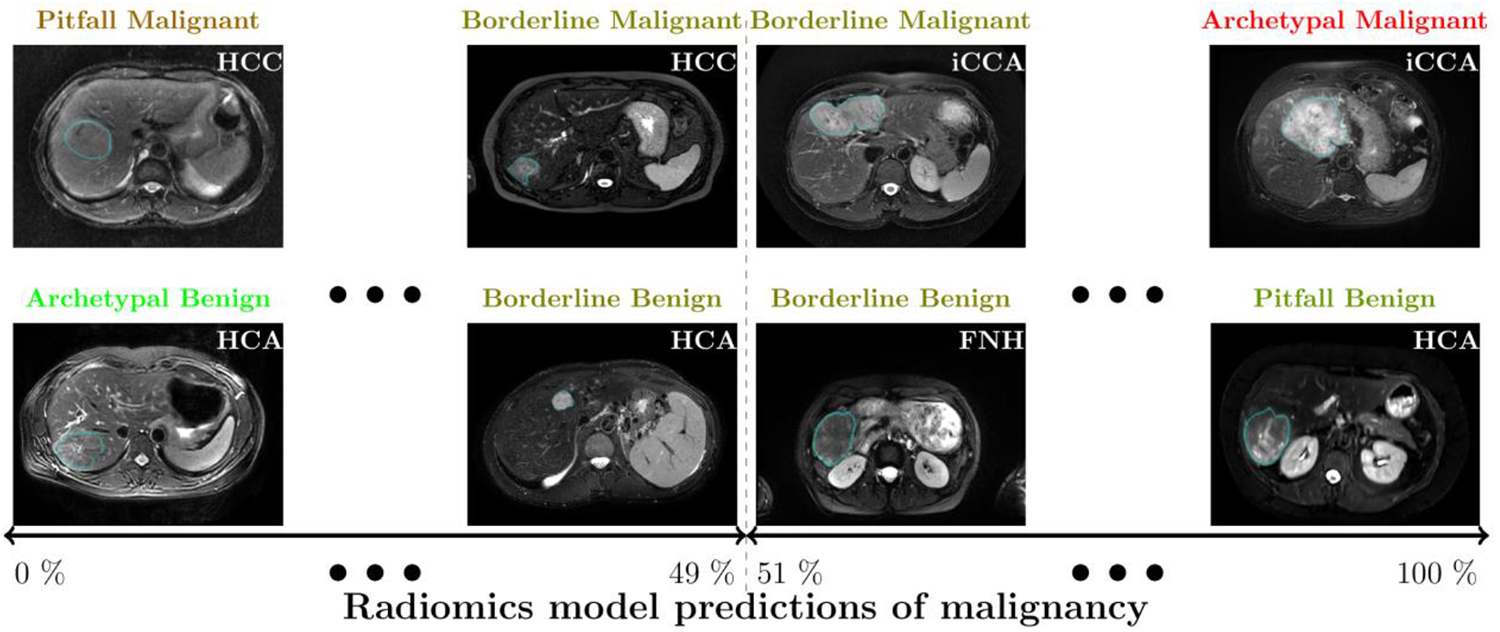
Examples of liver lesions on T2-weighted MRI. From left to right, examples of lesions considered by the radiomics model as archetypal (i.e., predicted probability close to extremes and correct), pitfall (i.e., predicted probability close to extremes and incorrect), and borderline (i.e., predicted probability close to border of 50%). Abbreviations: HCC: hepatocellular carcinoma; iCCA: intrahepatic cholangiocarcinoma; HCA: hepatocellular adenoma; FNH: focal nodular hyperplasia.

The predictions by the radiomics model on dataset C were compared to the characteristic scores of Radiologist 1, who had the highest performance. The correlation between the probability of malignancy as predicted by the radiomics model and heterogeneity as scored by Radiologist 1 was moderate (Pearson coefficient: 0.58). Radiologist 1 performed well when lesions had an apparent atoll sign: from the 19 lesions which Radiologist 1 scored as having an atoll sign and therefore classified as benign, 17 were indeed benign and 2 malignant. On the contrary, the radiomics model only classified 11 of these lesions correctly, but these included the 2 malignant lesions misclassified by Radiologist 1.

## Discussion

In this study, we developed a radiomics model to distinguish between malignant and benign primary solid liver lesions based on T2-weighted MRI in patients with non-cirrhotic livers. We showed that our radiomics model can distinguish between these lesions, both in an internal cross-validation and in two external validations.

The substantial increase of radiomics related research in recent years has led to various guidelines, vulnerabilities, and gaps (22-24, 42). While several studies have evaluated radiomics for the classification of liver lesions (16-18), radiomics for primary liver cancer is still in the early stages, and many of these aspects still need to be addressed (21). One of the most important is external validation, which is crucial to ensure a high level of evidence in a variety of settings (22, 23). Furthermore, the lack of standard imaging parameters can be problematic as these can affect the appearance of the lesion and thus radiomics (21, 25). Requiring a comprehensive, standardized set of multiple MRI sequences is hardly feasible in practice. In this study, we therefore only used T2-weighted MRI without strict protocol requirements, and externally validated our model on two multicenter cohorts from different countries to assess the generalizability. The scans of the 486 patients included in this study originated from 159 different MRI scanners, resulting in substantial heterogeneity in the acquisition protocols. In univariate analyses, only four radiomics features showed statistically significant differences in all three datasets. Nevertheless, our method performed well on data from unseen scanners (i.e., not present in the training dataset), indicating good generalizability. Furthermore, we used routinely acquired T2-weighted MRI, increasing the chance that the reported performance can be reproduced in a routine clinical setting. All lesions in our study, except typical FNH (27), were pathologically proven to ensure the ground truth was objective. We also set inclusion criteria to maximize the relevance to clinical decision making. Usage of a single, widely used sequence and the fact that the lesion phenotypes included in our study present more than 90% of all solid lesions, makes our model widely applicable.

To compare the radiomics model to routine clinical practice, the model’s predictions were compared to assessment by two experienced abdominal radiologists. The agreement between radiologists was moderate, indicating some observer variation in the predictions. The characteristics apparently used by the radiomics model to define lesions as archetypal, borderline, and pitfalls, were different than those used in the scoring of the radiologists. This is also illustrated by the moderate correlation in the heterogeneity scored by Radiologist 1 and the radiomics model’s score, and their different predictions on lesions with an apparent atoll sign. As these results indicate the potential complementary value of the radiomics model, further research should focus on how the radiologists’ and the radiomics model’s predictions can be optimally combined to improve clinical decision making.

Our results indicate that assessment of primary solid liver lesions by radiologists can be challenging and is subject to observer dependence. Existing guidelines may aid the radiologist in specific scenarios, such as EASL’s guidelines for management of benign liver tumors (4) and HCC (5), or LI-RADS for patients with cirrhotic livers (19). In this study, inclusion and exclusion criteria were determined to maximize the clinical relevance, covering scenarios not included in these guidelines. Our radiomics model therefore complements these existing initiatives. Radiomics may be especially useful on lesions where there is no consensus between radiologists, or on the pitfalls for radiologists. Additionally, it may serve as a gatekeeper in non-specialized centers, shortening the diagnostic delay by enabling direct referral to an expertise center and reducing the number of missed malignant lesions.

Age and sex are known to be strong predictors for distinguishing malignant from benign liver lesions (1, 26). In our study, in line with worldwide findings, (young) females represented the majority of benign lesions, while older patients represented the majority of malignant lesions (1, 26). The models based on age and sex used an age threshold at 49 years. In dataset C, only 19 (17%) of the 114 lesions of patients below 49 years were malignant. Although this therefore yielded a good overall performance, it would lead to missing all malignant lesions in young patients, for whom such a diagnosis is essential as these patients would benefit most from treatment. Simply classifying all lesions below 49 years as benign, regardless of any imaging information, would be unacceptable and cannot be applied to the general population. On the other hand, the radiomics model purely based on T2-weighted MRI does not use any population-based information. The model rather predicts the probability of a lesion being malignant based on the imaging appearance. Our radiomics method could be especially useful in young males to not miss malignant lesions, and in older females to detect benign lesions. Future research should therefore also focus on optimally combining imaging, age, and sex.

Our study has several limitations. First, while the inclusion and exclusion criteria were set to maximize the relevance to clinical decision making, they limit the applicability, as our model cannot be applied to all liver lesions, and may have led to selection biases. Future research should therefore focus on loosening these criteria, for example including patients with smaller lesions (maximum diameter < 3 cm), liver disease, more typical lesions, i.e., that are routinely not biopsied, and other (rare) phenotypes. Second, the current radiomics approach requires semi-automatic segmentations. While accurate, this process is time consuming and subject to some observer variability, limiting the transition to clinical practice. We do not believe that this has substantially affected the results, as the inter-observer DSC indicated good segmentation reproducibility, and the radiomics model performed similar in the internal and external validations despite training and testing on segmentations of various observers. Automatic segmentation methods, for example with deep learning (43), may help to further automate the method and avoid observer dependence.

On one hand, using a single, widely available (T2-weighted) MRI sequence without strict protocol restrictions is a strength of our model. On the other hand, in real life, radiologists use multiple sequences in their assessment, indicating that a multi-sequence model may lead to an improved performance. EASL’s guidelines also describe lesion assessment characteristics based on these other sequences, e.g. wash-out on dynamic contrast enhanced T1-weighted MRI, and diffusion restrictions (low ADCs) (4, 5). These other sequences may contain additional information to improve the radiomics and radiologists’ performance (16). Especially when extending our work to phenotyping, these sequences may contain essential information for an accurate diagnosis. Main additional challenges for such a multi-sequence model, due to the lack of a standardized protocol in the literature, are the additional heterogeneity, missing data as not all these sequences are acquired by default, and overcoming differences in appearance caused by the variations in contrast agents (44). We used only T2-weighted MRI, as this sequence suffers less from these disadvantages; is widely available, thus a T2-weighted MRI based radiomics model is feasible to use in routine clinical practice; is relatively simple and thus showing less heterogeneity as e.g. sequences with contrast; is reliable for lesion segmentation; and is minimally sensitive to motion or breathing artefacts; and is informative (4, 5, 19). The latter is also illustrated by our results, as the two radiologists were already able to distinguish malignant from benign lesions quite accurately using only T2-weighted MRI.

Future research should, besides the points mentioned in the previous paragraphs, focus on extending our work to phenotyping (e.g. HCC, iCCA, HCA, FNH), and possibly even subtyping (e.g. inflammatory HCA, β-catenin activated HCA) to further aid clinical decision making. Furthermore, to gain better insight into the complementary value of radiomics, our model may be compared with more radiologists. In our study, two experienced abdominal radiologists who were trained at the same center scored the patients. Hence, it would be valuable to compare with radiologists from a variety of institutes, also including less experienced and non-academic radiologists. This will also give a better insight into which type of lesions are difficult for radiologists to classify or reach consensus on, and thus where radiomics could have the highest added value.

In conclusion, our radiomics model based on T2-weighted MRI was able to distinguish malignant from benign primary solid liver lesions in patients with non-cirrhotic livers, both in an internal validation and in two external validations on heterogeneous, multicenter data. Pending further optimization and generalization, our model may serve as a robust, non-invasive and low-cost aid to enable quicker referral and refine patient selection prior to biopsies, and help solve the shortage of radiologists (45).

## Data Availability

Imaging and clinical research data are not available at this time. Programming code is available on Zenodo at DOI https://doi.org/10.5281/zenodo.5175705.

https://github.com/MStarmans91/LiverRadiomics

## Acknowledgments

Martijn Starmans acknowledges funding from the research program STRaTeGy (project number 14929-14930), which is (partly) financed by the Netherlands Organisation for Scientific Research (NWO). This work was partially carried out on the Dutch national e-infrastructure with the support of SURF Cooperative.

## Abbreviations

AUC: area under the curve
ADC: apparent diffusion coefficient
DSC: Dice similarity coefficient
EASL: European association for the study of the liver
FNH: focal nodular hyperplasia
HCA: hepatocellular adenoma
HCC: hepatocellular carcinoma
iCCA: intrahepatic cholangiocarcinoma
MRI: magnetic resonance imaging
ROC: receiver operating characteristic
WORC: workflow for optimal radiomics classification

