## Supplementary Materials for "Automated differentiation of malignant and benign primary solid liver lesions on MRI: an externally validated radiomics model"

** equal contributions*

**Supplementary Material 1: Pathological examination**

In the pathology, a distorted (micro)architecture of liver tissue was the common feature of the included lesions. Histomorphology often combined with (immuno)histochemistry served the final diagnosis. Hepatocellular lesions with loss of portal tracts, cell atypia, thick trabeculae (loss of reticulin fibers), pseudoglandular transformation, isolated small arterial branches, and capillarization of the sinusoidal areas (CD34 positive) with supportive immunohistochemistry (glypican-3, glutamine synthetase, HSP-70), were classified as HCC (1). Cases where the reticulin fibers were maintained, the pseudoglandular transformation and the cell atypia were absent or minimal, and the immunohistochemistry (glypican-3, HSP-70) was negative, were classified as HCA (1). Lesions composed of non-organoid arranged glandular structures, localized at the periphery of the second-order bile ducts with an expression of keratin 7 and 19, were classified as iCCA, either conventional or cholangiolocarcinoma (2). Non-neoplastic lesions, composed of hyperplastic hepatocellular nodules separated by fibrotic septa, creating a microscopic image of “localized cirrhosis” and often centrally a scar, were classified as FNH. Glutamine synthetase showed the pathognomonic “map-like” pattern of immunohistochemical expression (anastomosing groups of positively stained hepatocytes (3)).

**Supplementary Material 2: Radiomics feature extraction**

This supplementary material is similar to (4, 5), but details relevant for the current study are highlighted.

A total of 564 radiomics features were used in this study. All features were extracted using the defaults for MRI scans from the Workflow for Optimal Radiomics Classification (WORC) (6), which internally uses the PREDICT (7) and PyRadiomics (8) feature extraction toolboxes. An overview of all features is depicted in **Supplementary Table S2**. For details on the mathematical formulation of the features, we refer the reader (9). More details on the extracted features can be found in the documentation of the respective toolboxes, mainly the WORC documentation (10).

For MRI scans, the images are by default normalized in WORC as the scans do not have a fixed unit and scale, contrary to e.g. computed tomography (Hounsfield units). Normalization is performed using z-scoring, i.e., subtracting the mean and dividing by the standard deviation. As the datasets used in this study exhibit substantial heterogeneity in the acquisition protocols, the mean and standard deviation were computed based on the segmentation of the regions of interest (ROIs), i.e., the lesions, and not on the full image, as the latter is more sensitive to acquisition variations. The images were not resampled, as this would result in interpolation errors, especially in the axial direction due to the substantial differences in slice thicknesses. The code to extract the features has been published open-source (11).

The features can be divided in several groups. Thirteen intensity features were extracted using the histogram of all intensity values within the ROIs and included several first-order statistics such as the mean, standard deviation and kurtosis. These describe the distribution of intensities within the lesion. Thirty-five shape features were extracted based only on the ROI, i.e. not using the image, and included shape descriptions such as the volume, compactness and circular variance. These describe the morphological properties of the lesion. Nine orientation features were used, describing the orientation of the ROI, i.e. not using the image. Lastly, 507 texture features were extracted using Gabor filters (156 features) (9), Laplacian of Gaussian filters (39 features) (9), vessel (i.e. tubular structures) filters (39 features) (12), the Gray Level Co-occurrence Matrix (144 features) (9), the Gray Level Size Zone Matrix (16 features) (9), the Gray Level Run Length Matrix (16 features) (9), the Gray Level Dependence Matrix (14 features) (9), the Neighbourhood Grey Tone Difference Matrix (5 features) (9), Local Binary Patterns (39 features) (13), and Local Phase filters (39 features) (14, 15). These features describe more complex patterns within the lesion, such as heterogeneity, presence of blob-like structures, and presence of line patterns.

Most of the texture features include parameters to be set for the extraction. The values of the parameters that will result in features with the highest discriminative power for the classification at hand (i.e., malignant versus benign) are not known beforehand. Including these parameters in the workflow optimization, see **Supplementary Material 3**, would lead to repeated computation of the features, resulting in a redundant increase in computation time. Therefore, alternatively, these features are extracted at a range of parameters as is default in WORC. The hypothesis is that the features with high discriminative power will be selected by the feature selection methods and/or the machine learning methods as described in **Supplementary Material 3**. The parameters used are described in **Supplementary Table S2**.

The variations in the slice thickness due to the heterogeneity in the acquisition protocols may cause feature values to be dependent on the acquisition protocol. Moreover, the slice thickness is substantially larger than the pixel spacing. Hence, extracting robust 3D features may be hampered by these variations, especially for low resolutions. To overcome this issue, all features were extracted per 2D axial slice and aggregated over all slices, which is default in WORC. Afterwards, several first-order statistics over the feature distributions were evaluated and used in the machine learning approach.

**Supplementary Material 3: Radiomics decision model creation**

This appendix is similar to (4, 5), but details relevant for the current study are highlighted.

The Workflow for Optimal Radiomics Classification (WORC) toolbox (6) makes use of automated machine learning to create the optimal performing workflow from a variety of algorithms. Besides deciding whether to use an algorithm, most algorithms require hyperparameters, i.e., parameters that need to be set before the actual learning step, to be tuned to enhance the performance. WORC defines a workflow as a specific sequential combination of algorithms and their respective hyperparameters. In WORC, the radiomics workflow is split into the following components: image and segmentation preprocessing, feature extraction, feature and sample preprocessing, and machine learning. For each component, a collection of algorithms and their associated hyperparameters is included. Given this search space, WORC uses automated machine learning to find the optimal solution. The code to use WORC for creating the decision models in this specific study has been published open-source (11).

The workflows could be constructed from the following default search space in WORC, which components can only be combined in the order listed below:

1. Features selection: a group-wise search, in which specific groups of features (i.e., intensity, shape, and the subgroups of texture features as defined in **Supplemental Material 2** and **Supplementary Table S2**) are selected or deleted. To this end, each feature group had an on/off variable which is randomly activated or deactivated, which were all included as hyperparameters in the optimization.
2. Feature imputation: when a feature could not be computed, e.g. a lesion is too small for a specific feature to be extracted, a feature imputation algorithm is used to estimate replacement values for the missing values. Strategies for imputation included 1) the mean; 2) the median; 3) the mode; 4) a constant (default: zero); and 5) a nearest neighbor approach.

3. Feature selection: a variance threshold, in which features with a low variance (<0.01) are removed. This method was always used, as this serves as a feature sanity check with almost zero risk of removing relevant features.
4. Feature scaling was performed to make all features have the same scale, as otherwise the machine learning methods may focus only on those features with large values. This was done through z-scoring, i.e., subtracting the mean value followed by division by the standard deviation, for each individual feature. A robust version of z-scoring was used, in which outliers, i.e., values below the 5th percentile or above the 95th percentile, were excluded from computing the mean and variance.
5. Feature selection: optionally, the RELIEF method (16), which ranks the features according the differences between neighboring samples. Features with more differences between neighbors of different classes (i.e., malignant versus benign) are considered higher in rank.
6. Feature selection: optionally, features are selected by training a machine learning model and selecting features that are regarded important by the model. Hence the used model should be able to give the features an importance weight. Included model choices are LASSO, logistic regression, and a random forest.
7. Dimensionality reduction: optionally, principal component analysis (PCA) is used, in which either only those linear combinations of features were kept which explained 95% of the variance in the features or a limited number of components (between 10 – 50).
8. Feature selection: optionally, individual feature are selected through univariate testing. To this end, for each feature, a Mann-Whitney U test was performed to test for significant differences in distribution between the labels (i.e., malignant versus benign). Afterwards, only features with a p-value above a certain threshold were selected.
9. Resampling: optionally, a various resampling strategy could be used, which are used to overcome class imbalances and reduce overfitting on specific training samples. These included various methods from the imbalanced-learn toolbox (17): random over-sampling, random under-

sampling, near-miss resampling, the neighborhood cleaning rule, ADASYN, and SMOTE (regular, borderline, Tomek and the edited nearest neighbors variant).

10. Machine learning: lastly, a machine learning method is used to determine a decision rule to distinguish the classes. Methods included were; 1) logistic regression; 2) support vector machines; 3) random forests; 4) naive Bayes; 5) linear discriminant analysis; 6) quadratic discriminant analysis; 7) AdaBoost (18); and 8) extreme gradient boosting (19).

By default in WORC, all model construction and optimization was performed on the training set in order to prevent overfitting on the test dataset. To prevent overfitting on the *training* dataset, a 5x random-split stratified cross-validation (20, 21) was performed within the training dataset as well, using 85% for model training and 15% for model validation, see **Supplementary Figure S1**.

WORC states the radiomics workflow as a combined algorithm selection and hyperparameter optimization problem (CASH), as algorithm selection and hyperparameter optimization are often not independent (22). Within the training dataset, CASH optimization is performed by testing thousand pseudo-randomly generated radiomics workflows from the above search space. These are trained on the five training datasets in the 5x random-split training-validation cross-validation, and ranked according to their mean performance on the five validation datasets. As performance metric, the weighted F1-score is used, which is the harmonic average of the precision and recall.

Using only the single workflow that on average performs best on the validation datasets may result in poor generalization due to overfitting on the validation datasets. Hence, an ensemble was constructed by combining the workflows that perform best on the validation datasets. Ensembling was done using the default of WORC by averaging the posteriors of the 100 best workflows.

The following pseudo code illustrates the algorithm of WORC:

- **For** each 100x random-split training-test cross-validation iteration:
    - **Do:** Construct the training dataset by randomly selecting 80% of the patients.

- **Do:** On this training dataset, define 5x random-split cross-validation splits, selecting in each iteration 85% of the patients for training and 15% for validation.
- **Do:** Pseudo-randomly sample 1,000 workflows from the search space.
- **For** each of the 1,000 sampled workflows:
  - **Do:** Train the workflow on the five training datasets in the 5x random-split cross-validation.
  - **Do:** Compute the mean weighted F1-score on the corresponding five validation datasets in the 5x random-split cross-validation.
- **Do**: Rank the 1,000 workflows, retrain the best 100 workflows on the full training dataset and combine them in an ensemble.
- **Do:** Evaluate “the model”, i.e., the ensemble of the best 100 workflows as trained on the training dataset, on the test dataset, i.e., the remaining 20% of the patients that were not included in the training dataset.

The largest experiments in this study consists of executing 500,000 workflows (1,000 pseudo-randomly generated workflows, times a 5x train-validation cross-validation, times 100x train-test cross-validation for the internal validation), which can be parallelized. The computation time of training or testing a single workflow is on average less than a second, depending on the size of the dataset both in terms of samples (i.e. patients) and features. The largest experiment in this study, i.e. the internal validation on dataset A, had a computation time of approximately 24 hours on a 32 CPU core machine. The contribution of the feature extraction to the computation time was negligible.

The code for the radiomics feature extraction and model creation, including more details, has been published open-source (11).

### Supplementary Figures and Tables

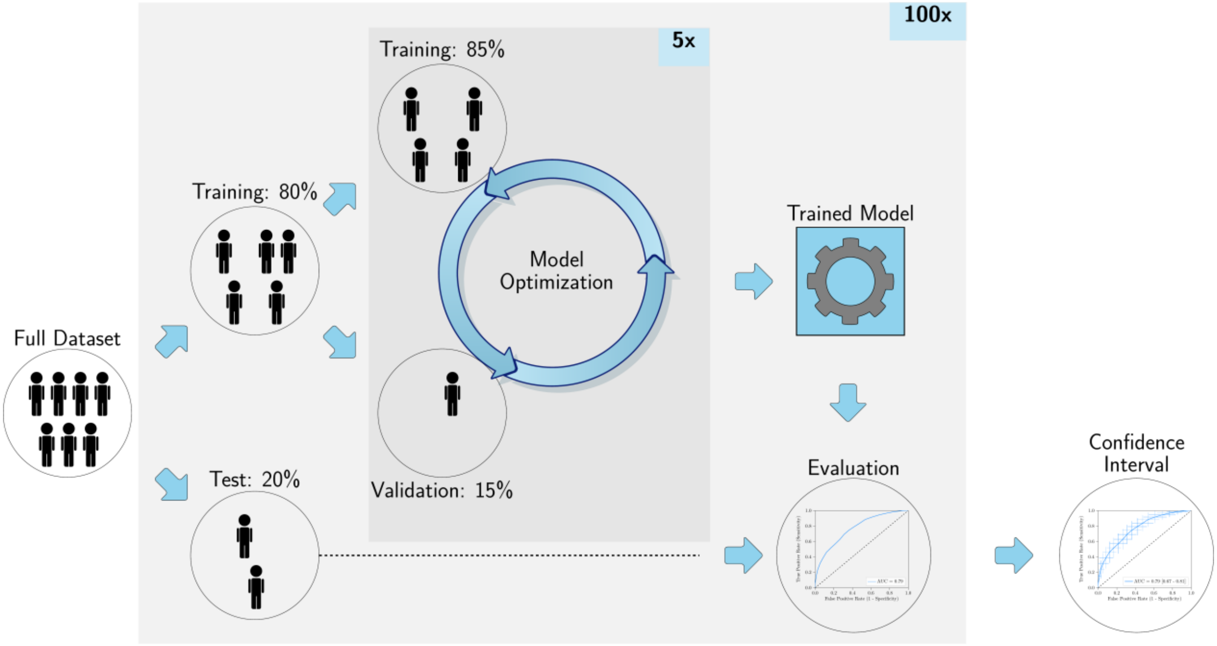

**A. Internal validation**

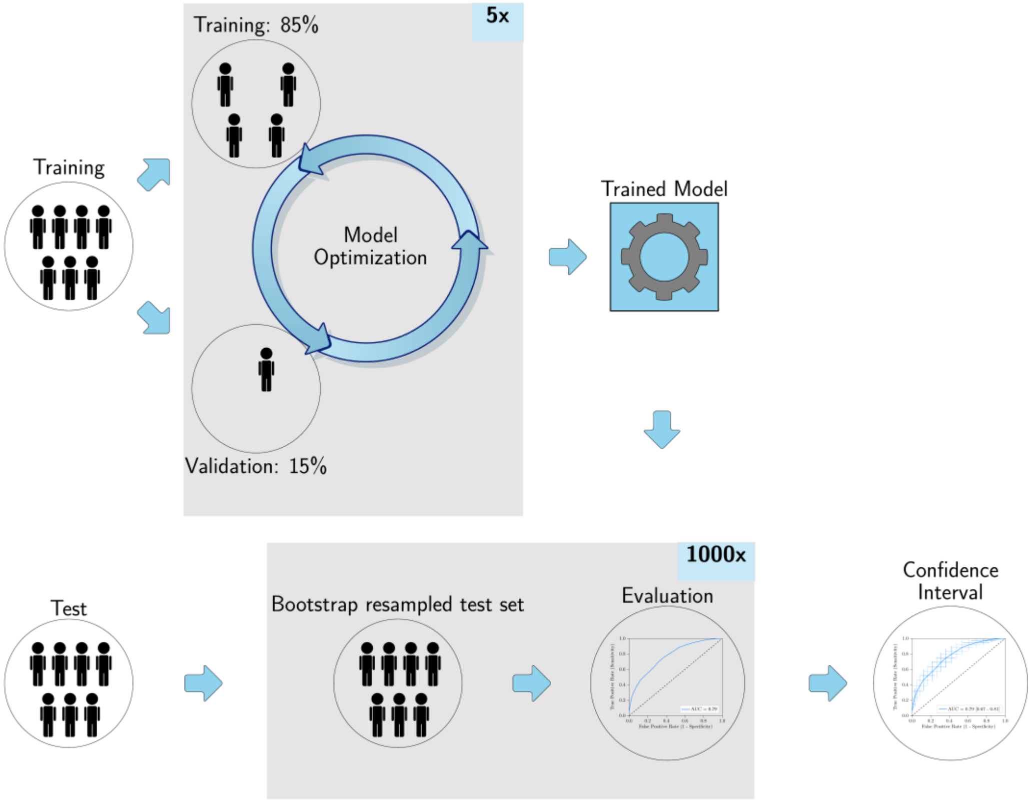

**B. External validation**

***Supplementary Figure S1. Visualization of evaluation setups***. *(A) The 100x random-split cross-validation used in the internal validation; (B) and the 1,000x bootstrap resampling in the external validations. Both include an internal random-split cross-validation within the training dataset for the model optimization.*

***Supplementary Table S1. Overview of univariate testing of radiomics feature****s. Per dataset (A, B, and C), the statistical significance of the difference between the malignant and benign lesions was assessed using a Mann-Whitney U test for continuous variables, and a Chi-square test for discrete variables. Only the features that showed statistically significant differences in dataset A are include. All p-values were corrected for multiple testing by multiplying the p-values with the total number of tests (564). Statistically significant p-values and names of that showed statistically significant differences in all three datasets are given in* ***bold****.*

| Feature name | p-value A | p-value B | p-value C |
|---|---|---|---|
| tf_kurtosis_sigma1 | **$9.26\times10^{-10}$** | **$1.80\times10^{-5}$** | 1.00 |
| tf_mean_sigma1 | **$1.06\times10^{-8}$** | **$1.27\times10^{-4}$** | 1.00 |
| tf_LBP_std_R3_P12 | **$3.19\times10^{-8}$** | **$8.60\times10^{-4}$** | 1.00 |
| **tf_LBP_quartile_range_R8_P24** | **$1.56\times10^{-7}$** | **0.0026** | **$7.77\times10^{-5}$** |
| tf_peak_sigma1 | **$2.74\times10^{-7}$** | **0.0028** | 1.00 |
| tf_median_sigma1 | **$8.27\times10^{-7}$** | **0.0035** | 1.00 |
| **tf_LBP_skewness_R8_P24** | **$1.33\times10^{-6}$** | **0,0067** | **$2{,}16\times10^{-4}$** |
| **tf_LBP_kurtosis_R8_P24** | **$1.51\times10^{-6}$** | **0.013** | **$9.44\times10^{-5}$** |
| **tf_LBP_mean_R8_P24** | **$1.53\times10^{-6}$** | **0.014** | **$2.20\times10^{-4}$** |
| tf_LBP_skewness_R15_P36 | **$5.18\times10^{-5}$** | 0.13 | **$8.70\times10^{-4}$** |
| tf_LBP_mean_R15_P36 | **$6.18\times10^{-5}$** | 0.14 | **$9.92\times10^{-4}$** |
| tf_mean_sigma10 | **$8.40\times10^{-5}$** | 0.94 | 1.00 |
| tf_LBP_kurtosis_R15_P36 | **$9.02\times10^{-5}$** | 0.19 | **$4.75\times10^{-4}$** |
| tf_LBP_median_R3_P12 | **$1.29\times10^{-4}$** | 0.086 | 0.27 |
| sf_area_min_2D | **$2.29\times10^{-4}$** | 0.67 | 1.00 |
| tf_Gabor_std_F0.2_A0.79 | **$3.65\times10^{-4}$** | **$6.42\times10^{-5}$** | 0.50 |
| tf_Gabor_kurtosis_F0.05_A0.79 | **$6.24\times10^{-4}$** | 1.00 | 1.00 |
| tf_LBP_skewness_R3_P12 | **$8.44\times10^{-4}$** | 0.28 | 0.17 |
| tf_Gabor_quartile_range_F0.2_A0.79 | **0.001** | **$9.71\times10^{-5}$** | 0.11 |
| tf_median_sigma10 | **0.001** | 0.51 | 1.00 |
| tf_Gabor_quartile_range_F0.2_A1.57 | **0.001** | **$3.79\times10^{-5}$** | 1.00 |
| tf_Gabor_max_F0.2_A0.79 | **0.004** | **$2.26\times10^{-4}$** | 0.24 |
| tf_Gabor_std_F0.2_A0.0 | **0.006** | **0.001** | 0.18 |
| tf_Gabor_std_F0.2_A1.57 | **0.008** | **0.002** | 1.00 |
| tf_Gabor_range_F0.2_A0.79 | **0.008** | **$6.92\times10^{-4}$** | 0.69 |
| tf_Gabor_quartile_range_F0.2_A2.36 | **0.009** | **$6.92\times10^{-4}$** | 0.12 |
| tf_LBP_mean_R3_P12 | **0.012** | 1.00 | 0.45 |
| tf_kurtosis_sigma10 | **0.012** | 0.73 | 1.00 |
| tf_std_sigma1 | **0.014** | 0.44 | 1.00 |
| tf_LBP_std_R15_P36 | **0.015** | 1.00 | 0.002 |
| tf_Gabor_max_F0.2_A1.57 | **0.015** | **0.001** | 1.00 |
| tf_Gabor_median_F0.5_A0.0 | **0.015** | 1.00 | 1.00 |
| sf_area_avg_2D | **0.015** | **0.010** | 1.00 |
| tf_Gabor_min_F0.2_A0.79 | **0.017** | **0.005** | 1.00 |
| tf_LBP_std_R8_P24 | **0.019** | 1.00 | **$8.80\times10^{-4}$** |
| tf_LBP_quartile_range_R15_P36 | **0.020** | 1.00 | 0.084 |
| tf_Gabor_quartile_range_F0.2_A0.0 | **0.020** | **$5.80\times10^{-4}$** | 0.090 |
| sf_area_max_2D | **0.023** | **0.011** | 1.00 |
| sf_shape_Flatness | **0.038** | 1.00 | 1.00 |

| | | | |
|---|---|---|---|
| of_COM_y | **0.038** | 0.69 | 1.00 |
| tf_Frangi_inner_energy_SR(1.0. 10.0)_SS2.0 | **0.042** | **0.033** | 1.00 |
| tf_GLDM_SmallDependenceHighGrayLevelEmphasis | **0.046** | **0.020** | 1.00 |
| tf_max_sigma10 | **0.046** | 1.00 | 1.00 |
| tf_Frangi_edge_energy_SR(1.0. 10.0)_SS2.0 | **0.049** | 0.081 | 1.00 |
| tf_Frangi_full_energy_SR(1.0. 10.0)_SS2.0 | **0.049** | 0.081 | 1.00 |

*Abbreviations: tf: texture feature; sf: shape features; of: orientation feature.

***Supplementary Table S2. Overview of the 564 radiomics features used in this study.*** *GLCM features were calculated in four different directions (0, 45, 90, 135 degrees) using 16 gray levels and pixel distances of 1 and 3. LBP features were calculated using the following three parameter combinations: 1 pixel radius and 8 neighbours, 2 pixel radius and 12 neighbours, and 3 pixel radius and 16 neighbours. Gabor features were calculated using three different frequencies (0.05, 0.2, 0.5) and four different angles (0, 45, 90, 135 degrees). LoG features were calculated using three different widths of the Gaussian (1, 5 and 10 pixels). Vessel features were calculated using the full mask, the edge, and the inner region. Local phase features were calculated on the monogenic phase, phase congruency and phase symmetry.*

| **Histogram (13 features)** | **LoG (13*3=39 features)** | **Vessel (12*3=39 features)** | **GLCM (MS) (6*3*4*2=144 features)** | **Gabor (13*4*3=156 features)** | **NGTDM (5 features)** | **LBP (13*3=39 features)** |
|---|---|---|---|---|---|---|
| **min** | min | min | contrast (normal, MS mean + std) | min | busyness | min |
| **max** | max | max | dissimilarity (normal, MS mean + std) | max | coarseness | max |
| **mean** | mean | mean | homogeneity (normal, MS mean + std) | mean | complexity | mean |
| **median** | median | median | angular second moment (ASM) (normal, MS mean + std) | median | contrast | median |
| **std** | std | std | energy (normal, MS mean + std) | std | strength | std |
| **skewness** | skewness | skewness | correlation (normal, MS mean + std) | skewness | | skewness |
| **kurtosis** | kurtosis | kurtosis | | kurtosis | | kurtosis |
| **peak** | peak | peak | | peak | | peak |
| **peak position** | peak position | peak position | | peak position | | peak position |
| **range** | range | range | | range | | range |
| **energy** | energy | energy | | energy | | energy |
| **quartile range** | quartile | quartile | | quartile range | | quartile range |
| **entropy** | entropy | entropy | | entropy | | entropy |

| **GLSZM (16 features)** | **GLRM (16 features)** | **GLDM (14 features)** | **Shape (35 features)** | **Orientation (9 features)** | **Local phase (13*3=39 features)** |
|---|---|---|---|---|---|
| **Gray Level Non Uniformity** | Gray Level Non Uniformity | Dependence Entropy | compactness (mean + std) | theta_x | min |
| **Gray Level Non Uniformity Normalized** | Gray Level Non Uniformity Normalized | Dependence Non-Uniformity | radial distance (mean + std) | theta_y | max |
| **Gray Level Variance** | Gray Level Variance | Dependence Non-Uniformity Normalized | roughness (mean + std) | theta_z | mean |
| **High Gray Level Zone Emphasis** | High Gray Level Run Emphasis | Dependence Variance | convexity (mean + std) | COM index x | median |
| **Large Area Emphasis** | Long Run Emphasis | Gray Level Non-Uniformity | circular variance (mean + std) | COM index y | std |
| **Large Area High Gray Level Emphasis** | Long Run High Gray Level Emphasis | Gray Level Variance | principal axes ratio (mean + std) | COM index z | skewness |
| **Large Area Low Gray Level Emphasis** | Long Run Low Gray Level Emphasis | High Gray Level Emphasis | elliptic variance (mean + std) | COM x | kurtosis |
| **Low Gray Level Zone Emphasis** | Low Gray Level Run Emphasis | Large Dependence Emphasis | solidity (mean + std) | COM y | peak |
| **SizeZoneNonUniformity** | RunEntropy | Large Dependence High Gray Level Emphasis | area (mean, std, min + max | COM z | peak position |
| **SizeZoneNonUniformityNormalized** | RunLengthNonUniformity | Large Dependence Low Gray Level Emphasis | volume (total, mesh, volume) | | range |
| **SmallAreaEmphasis** | RunLengthNonUniformityNormalized | Low Gray Level Emphasis | elongation | | energy |
| **SmallAreaHighGrayLevelEmphasis** | RunPercentage | Small Dependence Emphasis | flatness | | quartile |
| **SmallAreaLowGrayLevelEmphasis** | RunVariance | Small Dependence High Gray Level Emphasis | least axis length | | entropy |
| **ZoneEntropy** | ShortRunEmphasis | Small Dependence Low Gray Level Emphasis | major axis length | | |
| **ZonePercentage** | ShortRunHighGrayLevelEmphasis | | minor axis length | | |
| **ZoneVariance** | ShortRunLowGrayLevelEmphasis | | maximum diameter 3D | | |
| | | | maximum diameter 2D (rows, columns, slices) | | |
| | | | sphericity | | |
| | | | surface area | | |
| | | | surface volume ratio | | |

*Abbreviations: COM: center of mass; GLCM: gray level co-occurrence matrix; MS: multi slice; NGTDM: neighborhood gray tone difference matrix; GLSZM: gray level size zone matrix; GLRLM: gray level run length matrix; LBP: local binary patterns; LoG: Laplacian of Gaussian; std: standard deviation.
